# Public Search Interests related to COVID-19: Insights from Google Search Trends in Bangladesh

**DOI:** 10.1101/2020.12.26.20248858

**Authors:** Mazbahul G. Ahamad, Monir U. Ahmed, Daniel R. Uden

## Abstract

**Objective:** Public response monitoring is critical to reducing COVID-19 infections and developing effective public health strategies. This study explored Google search trends to understand public responses to COVID-19 concerns in Bangladesh.

**Methods:** We used country-level Google search trends data to examine the association between Google search terms related to COVID-19 deaths, face masks, and COVID-19 vaccines and the actual and one-week lagged actual COVID-19 death counts from February 2, 2020, to December 19, 2020, in Bangladesh. Results: Search terms related to COVID-19 deaths, face masks, and COVID-19 vaccines increased and peaked during March and April, but then began declining gradually after June 2020. The mean relative search volume for face masks (35 points) was higher than for death information (8 points) and vaccines (16 points) throughout the study period, and searching for masks peaked (100 points) during the third week of March. Search interests for death information and face masks were negatively correlated with the actual and one-week lagged actual COVID-19 death counts.

**Conclusion:** In response to declining trends in COVID-19-related google search terms, policymakers should strengthen ongoing risk communication and preventive information dissemination programs to control and prevent COVID-19 cases and deaths.

## 1. Introduction

Globally, there are approximately 74.30 million confirmed COVID-19 cases and more than 1.67 million confirmed deaths as of December 19, 2020 [1]. To minimize infection and mortality rates, countries like Bangladesh have adopted various mitigation strategies, including wearing face masks [2]. In light of these public health guidelines, people are searching for information on current COVID-19 death statistics, prevention measures, such as facemasks, and vaccine-related information, all of which can be understood by analyzing Google search trends. Previous studies have identified Google searches as a monitoring tool to understand and predict people’s search [3–7]. Monitoring search trends in near real-time allows policymakers to modify and update existing public health strategies and create effective policies that influence people’s preventive behaviors. This study analyzed Google search trends in Bangladesh for terms related to COVID-19 deaths, face masks, and COVID-19 vaccines to determine whether these terms are associated with actual COVID-19 death counts and one-week lagged actual COVID-19 death counts. This approach and findings may be used as a public search-interest monitoring tool for customizing ongoing COVID-19-related risk communication and preventive information dissemination programs.

## 2. Methods

We explored the relative search volume (RSV) of Google search trends [8] related to the current impact of COVID-19 (i.e., death counts presented on www.worldometers.info), preventive measures (i.e., wearing face masks), and future solutions (i.e., a COVID-19 vaccine) between February 2, 2020, and December 19, 2020, in Bangladesh. The Google trends RSV indicates search interests for the selected location and time, ranging from 0 to 100 in value points. The highest value, 100, indicates the most frequently searched topic at a given time. We then compared the weekly RSVs of search trends with weekly actual COVID-19 death counts [9] to detect correlations (Pearson’s correlation coefficient) between searches related to death information (collected from the website: worldometers), a preventive behavior (mask), and a potential remedy (vaccine). We also estimated time-lagged correlation to determine if actual death counts in the previous week were correlated with “worldometers”, “mask”, and “vaccine” search volumes.

## 3. Results and discussion

Search terms related to COVID-19 death information from “worldometers” (linked to www.worldometers.info) were low at the beginning of the first coronavirus wave in early March and peaked (RSV: 100 points) at the end of the third week of April (Figure 1). During the study period, the mean relative search volume related to “worldometers” (8 points) was lower than searches regarding face “mask” (35 points) and COVID-19 “vaccine” (16 points). Searches for death count information were negatively correlated with actual death counts (r = −0.46, P = 0.00) and the one-week lagged actual death counts (r = −0.54, P = 0.00). Figure 2 shows the relationships between the actual and one-week lagged death counts and Google search terms. These findings diverge from general expectations and the results of several prior studies [3,6,10], although they are consistent with Lin et al. (2020). It is possible that the high early volume of Google search terms in March and April was l driven by panic searches spurred by news of a new global pandemic. In subsequent months, internet searches regarding COVID-19 may have decreased as a result of reports of a significantly lower COVID-19 death rate in Bangladesh than the United States and other countries [11].

**Figure 1.**
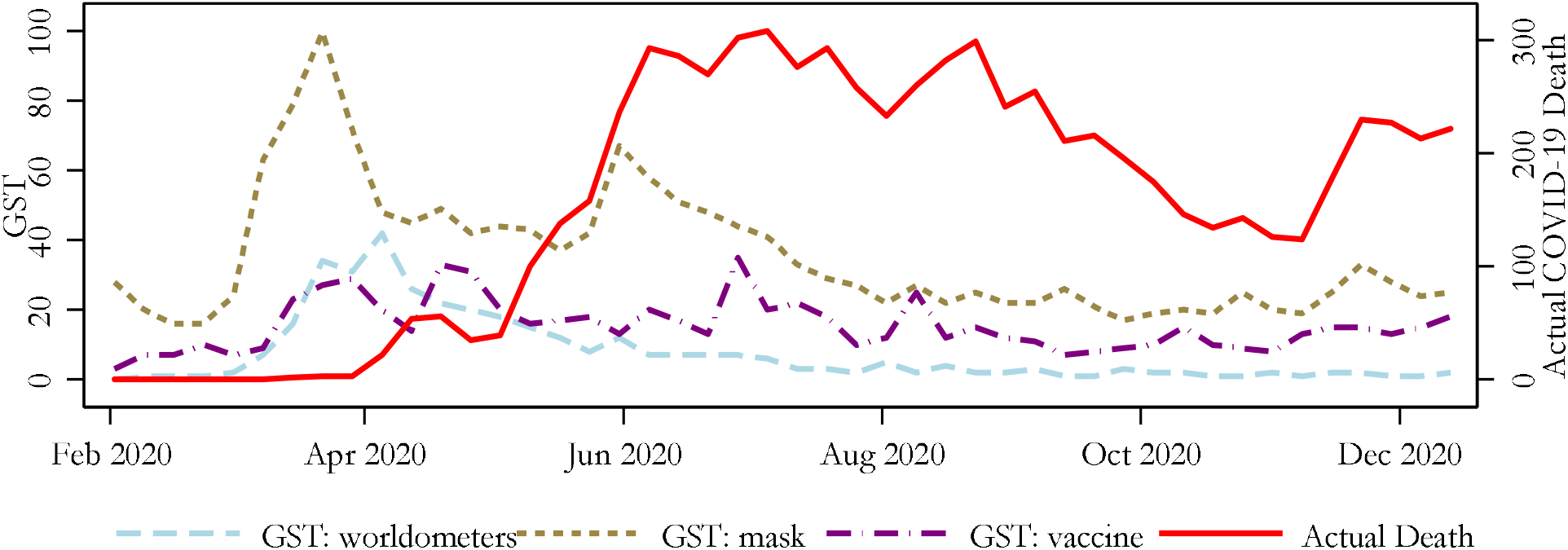
Google search trends on COVID-19 death counts, face masks, and vaccines (primary axis) and actual COVID-19 death counts (secondary axis), February–December 2020, Bangladesh.

**Figure 2.**
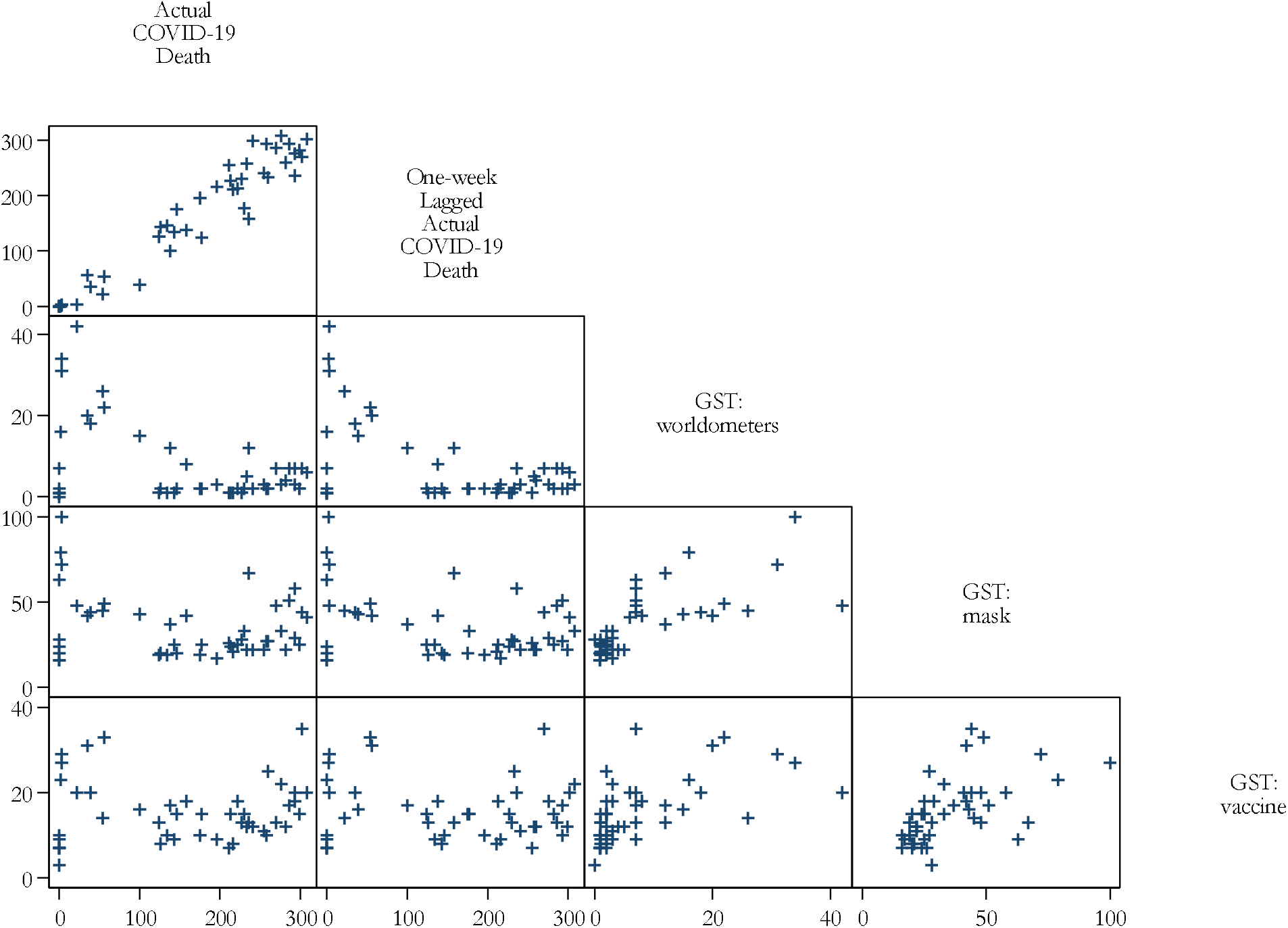
Scatter plot of actual COVID-19 death counts, one week-lag of actual COVID-19 death counts, and Google search terms related to COVID-19 deaths, face masks, and vaccines, February–December 2020, Bangladesh.

Mean relative search volume for face “mask” was greater than those for “worldometers” and “vaccine” throughout the study period (Figure 1)—reaching 100 points during the third week of March. After a downward trend, search interest for face “mask” reached 67 points again during the fourth week of May. Searches for “mask” were negatively correlated with actual death counts (r = −0.25, P = 0.09) and the one-week lagged actual death counts (r = −0.34, P = 0.02). This high relative search volume of mask was likely related to the government’s decision that people wear face masks in public places, which inspired people to purchase face masks online due to mask shortages at the beginning of April. Recently, most people in Bangladesh have neglected to wear masks, either due to the very low mortality rate or reduced government information dissemination and monitoring activities [12].

The mean relative search volume for COVID-19 “vaccine” was greater than that of COVID-19 death information (i.e., worldometers) but lower than that of for face “mask”. Both actual (r = −0.03, P = 0.83) and the one-week lagged actual death count (r = −0.08, P = 0.63) were not significantly correlated with search interest in a COVID-19 “vaccine”. However, search interest in a “vaccine” has increased in recent weeks (Figure 1), which may be explained by the recent approvals and availability of vaccines abroad during the second and third COVID-19 waves.

Google search trends for face masks is declining despite the actual death rate rising in recent weeks in Bangladesh. This decline in search-interests suggests that people may be ignoring guidelines to wear face masks, either due to the very low mortality rate in Bangladesh or moderate public health information dissemination and monitoring activities, which could cause more deaths in the second wave compared to the first wave. To minimize new infections and deaths, public health policymakers should use these Google search trends to help prioritize dissemination of additional COVID-19 prevention information, such as the wearing of face masks.

## Data Availability

The data are publicly available.

## Ethical approval

Ethical approval is not required for the study using publicly available data.

## References

[1] WHO. WHO Coronavirus Disease (COVID-19) Dashboard 2020. https://covid19.who.int/ (accessed December 16, 2020).

[2] Masrur A, Yu M, Luo W, Dewan A. Space-time patterns, change, and propagation of covid-19 risk relative to the intervention scenarios in Bangladesh. Int J Environ Res Public Health 2020;17:1–22.

[3] Husnayain A, Fuad A, Su E. Applications of Google Search Trends for risk communication in infectious disease management: A case study of the COVID-19 outbreak in Taiwan. Int J Infect Dis 2020;95:221–3.

[4] Lin Y, Liu C, Chiu Y. Google searches for the keywords of “wash hands” predict the speed of national spread of COVID-19 outbreak among 21 countries. Brain Behav Immun 2020;87:30–2.

[5] Gan Y, Jiang H, Li L, Yang Y, Wang C, Liu J, et al. Prevalence of burnout and associated factors among general practitioners in Hubei, China: A cross-sectional study. BMC Public Health 2019;19.

[6] Li C, Chen L, Chen X, Zhang M, Pang C, Chen H. Retrospective analysis of the possibility of predicting the COVID-19 outbreak from Internet searches and social media data, China, 2020. Eurosurveillance 2020;25:ppii=2000199.

[7] Szmuda T, Ali S, Hetzger T, Rosvall P, Słoniewski P. Are online searches for the novel coronavirus (COVID-19) related to media or epidemiology? A cross-sectional study. Int J Infect Dis 2020;97:386– 90. https://doi.org/10.1016/j.ijid.2020.06.028.

[8] Google Trends 2020. https://trends.google.com (accessed December 19, 2020).

[9] JHCRC. Johns Hopkins Coronavirus Resource Center 2020. https://coronavirus.jhu.edu/ x(accessed December 21, 2020).

[10] Effenberger M, Kronbichler A, Shin J, Mayer G, Tilg H, Perco P. Association of the COVID-19 pandemic with internet search volumes: a Google TrendsTM analysis. Int J Infect Dis 2020;95:192–7.

[11] Ahamad M, Tanin F, Talukder B, Ahmed M. Confirmed and ureported COVID-19-like illness death counts: an assessment of reporting discrepancy. Am J Trop Med Hyg 2020.

[12] Das T, Rahman M. Mandatory mask use mostly ignored across Bangladesh. New Age 2020. https://www.newagebd.net/article/120236/mandatory-mask-use-mostly-ignored-across-bangladesh x(accessed December 19, 2020).

